# Can objective measures of physical activity and sleep serve as early predictors of dementia among individuals with cognitive impairment?

**DOI:** 10.64898/2025.12.12.25342162

**Authors:** Victoria L. Orr, Karl Brown, Katie Wilson, Pamela Dean, Shu-Ching Hu, Daniel Levendowski, Edmund Seto, Andrew Shutes-David, Sarah Payne, Yeilim Cho, Debby W. Tsuang

**Author notes:** Dr. Dean is now affiliated with the Cleveland Clinic, OH, USA. Corresponding author: Debby W. Tsuang, MD, MSc; S182 GRECC, VA Puget Sound Health Care System, 1660 S. Columbian Way, Seattle, WA.

## Abstract

**Background and Objectives:** Physical activity and sleep are potential modifiable risk factors for the development of Alzheimer’s disease and related disorders (ADRD), but few studies have objectively measured both domains in participants across the cognitive continuum.

**Research Design and Methods:** Standard clinical assessment, accelerometry, and at-home EEG sleep data were obtained from older controls (n=9) and adults who met consensus diagnostic criteria for mild cognitive impairment (MCI; n=7), Alzheimer’s disease (AD; n=10), and dementia with Lewy bodies (DLB; n=11). Given these sample sizes, descriptive statistics are presented rather than formal statistical testing.

**Results:** The MCI group had the most surprising findings—although they were cognitively similar to the control group, they were less physically active than the AD group and had the worst sleep efficiency. The DLB group had the most severe motor and neuropsychiatric symptoms, were the least physically active, spent the least amount of time in rapid eye movement (REM) sleep, and spent the highest amount of time in non-REM sleep with hypotonia (NRH). The AD group had physical activity counts that fell between the DLB and control groups; REM sleep and NRH levels that were similar to the control and MCI groups; and autonomic activation index (AAI) and sleep spindle durations that were higher than the MCI and DLB groups.

**Discussion and Implications:** These findings highlight interesting physical activity and sleep patterns between groups, but larger samples are needed to investigate how objectively measured physical activity and sleep might serve as disease-specific digital biomarkers of neurodegenerative disorders.

**Translational Significance:** This study uses wearable technologies to measure physical activity and sleep in adults with and without cognitive impairment. The study found that adults with mild cognitive impairment had physical activity and sleep patterns that resembled people with dementia despite having cognitive scores that were closer to cognitively healthy controls. Sleep and activity patterns were distinct when comparing participants with Alzheimer’s and dementia with Lewy bodies. Larger studies are needed to validate these findings, but mobile health devices may be an accessible way to detect early cognitive decline and help differentiate dementia subtypes, resulting in earlier, targeted clinical care.

## BACKGROUND AND OBJECTIVES

Although no cure exists for Alzheimer’s disease, the recent approval of novel anti-amyloid therapies for Alzheimer’s disease (AD) may signal the dawning of a new era for the treatment of dementia.^1^ An important obstacle, however, is that many Americans with dementia or mild cognitive impairment (MCI) remain misdiagnosed or undiagnosed.^2,3^ These diagnostic delays increase caregiver and institutional burden while also prevent individuals from receiving treatments during the early stages of disease, which is when existing interventions are most likely to be effective.

Advances in neuroimaging, cerebrospinal fluid, and blood-based biomarkers can support the identification of AD and DLB, but these evaluation tools are invasive, costly, and primarily available in tertiary care settings, which limits their broader clinical applicability. To facilitate the improved identification of AD and related dementias (ADRD), including dementia with Lewy bodies (DLB), and to facilitate the widespread deployment of interventions, it will be necessary to develop less invasive and more scalable approaches.

In this study, we present preliminary efforts to leverage at-home digital technologies to detect physical and sleep-related features that may be indicative of early ADRD. Specifically, we aim to explore objective measures of physical activity and sleep, two physiological behaviors that may serve as early biomarkers and that have been characterized by some researchers as modifiable neuroprotective factors for the development of cognitive impairment in individuals with MCI and AD.^4,5^ We are particularly interested in accelerometry and at-home polysomnography (PSG), as these are objective, continuous measurements that can be obtained from adults in between clinic visits (e.g., in free-living conditions) and that can complement or enhance subjective self-report measures.^6,7^ Moreover, we hypothesize that these objective measures may be sensitive to the disturbances in motor function (e.g., parkinsonian symptoms, tremors, gait, and overall activity) and sleep (e.g., rapid eye movement [REM] behavior disorder [RBD]) that are hallmark features of DLB.^8^

Accelerometry studies have found, for instance, that prolonged sedentary activities may be associated with global cognitive decline and all-cause dementia.^9,10^ Indeed, individuals with cognitive impairments tend to be more sedentary and to engage in lower-intensity activity than individuals without cognitive impairment.^11,12^ In a longitudinal accelerometry study, Buchman et al.^9^ found that older adults with low daily physical activity had a 2.3 times greater risk of developing AD than older adults with higher activity levels.

PSG, which includes measurement of electroencephalography (EEG), is the gold standard for evaluating sleep,^13^ but traditional PSG is performed in hospital-based laboratories, where individuals often report a first-night effect of disrupted sleep.^14^Therefore, PSG/EEG devices like the Sleep Profiler have been developed to allow for continuous monitoring of sleep architecture in non-laboratory settings. Like accelerometry devices, these PSG devices are well tolerated by individuals with dementia.^15,16^ They have also been used to identify sleep disturbances, such as RBD.^17^

Despite the advancements in physical activity and sleep monitoring technology, few studies have used both kinds of mobile health devices to simultaneously measure sleep and physical activity in older adults with and without dementia. In a systematic review of sleep biomarkers in AD, for instance, Carpi et al.^18^ found that only one study utilized both accelerometry and EEG.

In addition, it may be useful to examine physical activity and sleep across the cognitive continuum, including in participants with no cognitive impairment, participants with early cognitive impairment, and participants with neurodegenerative disorders. Yet systematic reviews^19,20^ suggest that most studies focus on a single disorder or categorize participants as having dementia without specifically reporting the kind or severity. One key exception is McArdle et al.,^21^ who, like our earlier pilot study,^15^ suggested potential differences between individuals with AD and DLB in physical activity but found no statistically significant differences.

In our current study, we aim to address these gaps by providing initial characterizations of cognitively intact older adults as well as those with MCI, AD, and DLB based on data collected during the initial phase of our investigation.^15^ By integrating accelerometry and Sleep Profiler technology with cognitive and motor assessments in individuals across the neurocognitive spectrum, we aim to identify activity and sleep patterns that may help advance studies that aim to differentiate neurodegenerative disorders.

## RESEARCH DESIGN AND METHODS

### Participants

Individuals with no cognitive impairment (i.e., controls; n=9), MCI (n=7), AD (n=10), or DLB (n=11) were recruited from the VA Puget Sound Health Care System and the Puget Sound community under the approval of VA Puget Sound Institutional Review Board #1. All participants completed at least 8 years of education, were age 55 or older at enrollment, met a cognitive threshold per recent clinical records or study testing (i.e., 12 or higher on the Montreal Cognitive Assessment [MoCA] or 9 or higher on the telephone MoCA [t-MoCA]), were sufficiently ambulatory to measure physical activity using the accelerometry device, and had a family member, friend, or caregiver who was willing to answer questions about the participants’ cognition and functioning. Individuals were excluded if they had unstable psychiatric conditions, were regularly taking prescription-strength medications that were known to interfere with the Sleep Profiler (e.g., benzodiazepines and opioids), or had cognitive impairments not related to AD or DLB (e.g., stroke or head trauma). Control participants showed no cognitive impairment on the Modified Telephone Interview for Cognitive Status (TICS-M)^22^ and demonstrated no functional impairment; participants with MCI had a clinical diagnosis of MCI prior to joining the study, demonstrated cognitive impairment on the TICS-M, and/or described the presence of subtle functional impairment in participant and informant interviews; participants with AD met the National Institute on Aging-Alzheimer’s Association Workgroup criteria for probable AD;^23^ and participants with DLB met the Fourth Consensus Report criteria for probable DLB.^24^ After the initial study visit, all participant diagnostic designations were reviewed by a consensus conference of neurologists, psychiatrists, and study team members.

### Procedures

At the baseline visit, all participants completed a neuropsychological assessment that included the Montreal Cognitive Assessment (MoCA), the Repeatable Battery for the Assessment of Neuropsychological Status (RBANS), the Neuropsychiatric Inventory Questionnaire (NPI-Q), and Part III of the Movement Disorder Society (MDS)-modified Unified Parkinson’s Disease Rating Scale (UPDRS). Accelerometry and Sleep Profiler data were collected on consecutive days following the neuropsychological assessment.

#### MoCA

The MoCA version 7.1 is widely used in clinical and research settings as a screening tool for cognitive decline.^25^ It assesses short-term memory, visuospatial skills, executive function, attention, concentration and working memory, language, and orientation.

#### RBANS

The RBANS version A is a common clinical and research tool for characterizing mild impairment and dementia subtypes in geriatric populations.^26^ It includes five cognitive indexes—

#### MDS UPDRS Part III

The UPDRS is the standard assessment for the severity of motor symptoms relating to Parkinson’s disease and DLB.^27^ Part III includes 18 items that are rated on a scale of 0 (normal) to 4 (severe), including, for example, rigidity, bradykinesia, tremor, and postural/gait disturbance. In this study, Part III was performed by a qualified movement disorders neurologist.

#### NPI-Q

The NPI-Q is frequently used to assess neuropsychiatric symptoms in individuals with AD or other dementias by obtaining the perspectives of their caregivers.^28^ It evaluates 12 neuropsychiatric symptoms: delusions, hallucinations, agitation/aggression, depression/dysphoria, anxiety, euphoria/elation, apathy/indifference, disinhibition, irritability/lability, aberrant motor behavior, sleep disturbances, and appetite/eating changes. For each endorsed symptom, caregivers are asked to rate the frequency (1=rarely to 4=very often), severity (1=mild to 3=severe), and level of distress (0=not at all to 5=very severe). Here, we report binary *yes* or *no* responses to the neuropsychiatric symptoms of delusions and hallucinations.

#### Accelerometry

Participants wore triaxial wGT3X-BT accelerometry devices (ActiGraph/Ametris, Pensacola, FL) for 14 consecutive days and nights to collect activity data in the form of physical activity counts. The devices were deployed in person at the research visits, and participants wore the devices on the wrists of their nondominant hands. The start times were set to midnight of the day following the research visits to ensure a full 24 hours of data collection. Unless noted otherwise, the devices were worn all day, including when participants bathed or swam. The devices were set to a sampling rate of 30 Hz and were initialized using ActiLife software version 6.13.5. The accelerometry data were analyzed for all participants who had 10 or more days of data.

Daily activity counts quantify the overall amount of physical movement (and not only the number of steps) across a 24-hour period. Accelerometers continuously record movement intensity, which is converted into activity counts representing the sum of all detected accelerations over the course of the day. Higher daily activity counts indicate greater overall physical activity levels.

#### Sleep Monitoring

Participants wore the Sleep Profiler (Advanced Brain Monitoring, Inc., Carlsbad, CA) to collect sleep-related data. The device was affixed to the forehead with three sensors that acquire EEG, left and right electroocular activity, head movement, head position, pulse rate, and snoring sounds. Additional sensors were affixed to the chin and forearm to measure electromyographic activity. Participants with AD or DLB were asked to wear the Sleep Profilers for 2 consecutive nights. Controls or participants with MCI were added in the second phase of data collection, and for these participants, we modified the protocol to record 3 consecutive nights of Sleep Profiler data. Recordings with a minimum of four hours of recording time were considered for analysis.

Sleep variables included total REM sleep, non-REM sleep with hypertonia (NRH), sleep efficiency, autonomic activation index (AAI), sleep spindle duration, and atypical N3 (AN3). NRH is a novel biomarker that indicates increased muscle tone during non-REM sleep and is characteristic of parkinsonian spectrum disorders.^34^ Sleep efficiency is the ratio of actual sleep time to total time spent in bed, expressed as a percentage. Autonomic activation is based on the detection of a ≥6 beat-per-minute (BPM) increase and/or decrease compared to the previous 10th second, and the novel sleep biomarker autonomic activation index (AAI) reflects the number of these autonomic activation events detected per hour of sleep. Sleep spindles were auto detected based on temporal excursions in the absolute and relative sigma power in combination with simultaneous bursts of alpha power of at least 250 milliseconds; during these sigma and alpha power bursts, beta and electromyography power were tested to ensure relative suppression. Spindle lengths were restricted to 0.5 to 3.0 seconds in length, and spindle duration reflects the sum of all spindle lengths. Atypical N3 is slow-wave sleep (or “N3”) with abnormal electroencephalography patterns that do not fully match the classic definition of N3 per the American Academy of Sleep Medicine. Values for these variables of interest were averaged across nights with weighting based on the total sleep time per night.

### Statistical analysis

Descriptive statistics were calculated for all variables of interest. For continuous variables, we computed means and standard deviations. For categorical variables, we calculated frequencies and proportions. The number of participants with missing data was reported for each variable. All analyses were performed using R statistical software (version 4.5.1).^29^ Due to the planned small sample sizes for each group, we did not conduct formal statistical analysis across groups.

## RESULTS

Demographic and clinical characteristics of the various diagnostic groups are presented in Table 1 for all participants who met our inclusion and exclusion criteria. Given that the primary recruitment site was a Veterans hospital, most of the study participants were male and/or white, although the MCI group was more racially diverse. Participants with DLB had the lowest mean age, whereas participants with MCI or AD had the highest mean age.

**Table 1:**
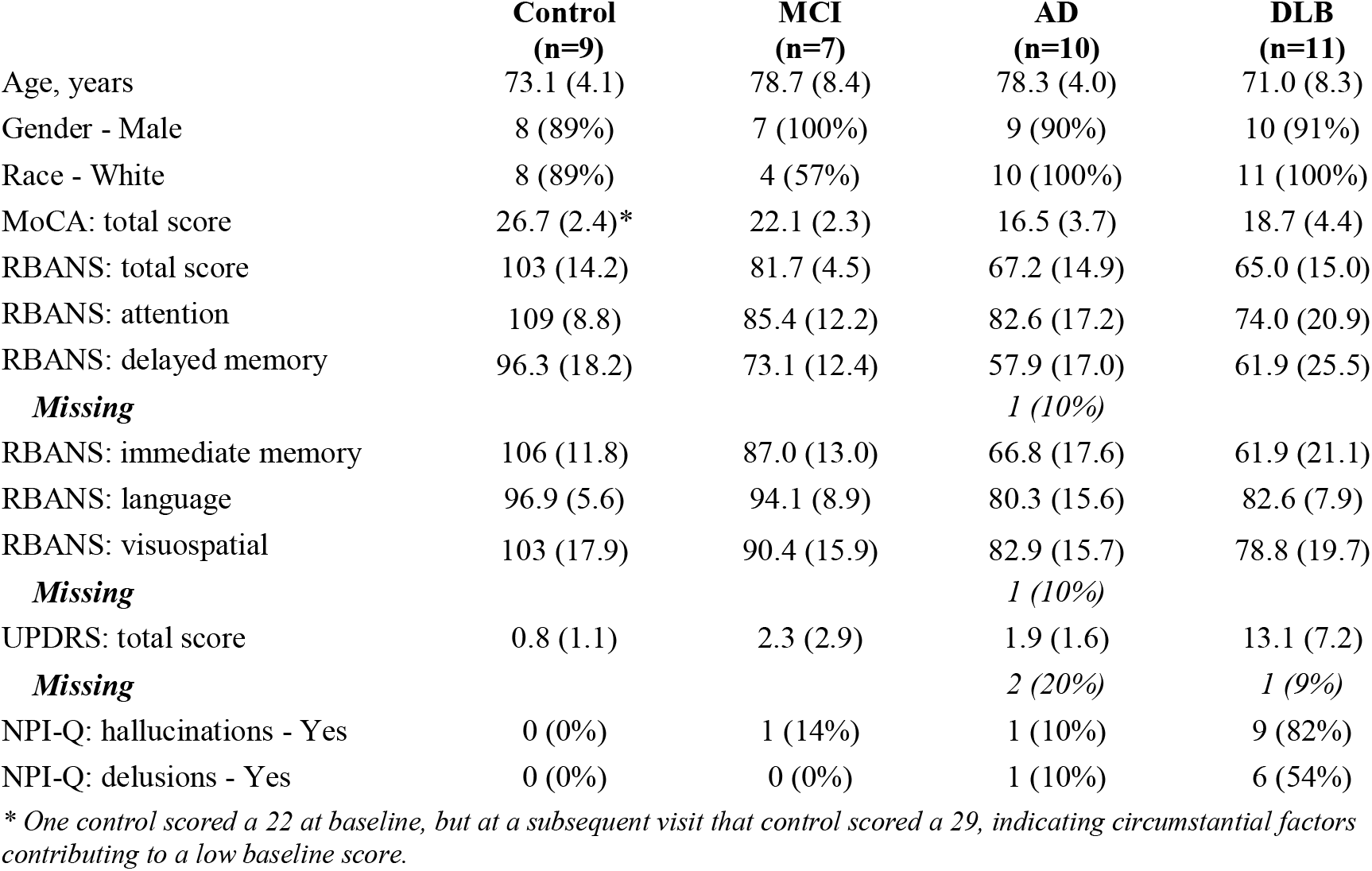
Demographic and clinical characteristics by diagnosis group.

The standard MoCA scoring paradigm characterizes individuals as having normal cognition (26+), mild cognitive impairment (18–25), moderate cognitive impairment (10–17), or severe impairment (<10).^30^ As shown in Table 1, our control group had a mean score in the normal cognition range, our MCI group had a mean score in the middle of the MCI range, our AD group had a mean score in the moderate cognitive impairment range, and our DLB group had a mean score at the lower end of the MCI range.

The RBANS total scores and domain scores were also as expected. As shown in Table 1, our control group had a mean total score that indicated average cognition, our MCI group had a mean total score that indicated low to average cognition, and our AD or DLB groups had mean total scores that fell in the impaired cognition range. The RBANS domain scores were similar, except that the participants with AD or DLB fell within the borderline impaired range (1.5–2.0 SD below mean) or the low-to-average range for the attention, language, and visuospatial domains.

On the MDS UPDRS Part III, the DLB group had markedly higher scores than the other groups; this is indicative of more severe parkinsonian motor symptoms and is consistent with the motor features commonly associated with this condition.

The NPI-Q also revealed differences in the DLB group compared to the other groups, as more than half of the DLB group reported hallucinations or delusions, whereas these symptoms were rare or nonexistent in the control, MCI, or AD groups.

Summaries of the digital biomarker variables measured in this study are shown in Table 2. In the context of physical activity, the DLB group was the least physically active, whereas the control group was the most physically active, and the MCI and AD groups exhibited intermediate activity levels between the DLB and control groups.

**Table 2:** Digital biomarker values by diagnosis group.

|  | **Control (n=9)** | **MCI (n=7)** | **AD (n=10)** | **DLB (n=11)** |
| --- | --- | --- | --- | --- |
| Daily activity counts (millions) | 2.2 (0.3) | 1.7 (0.4) | 1.6 (0.2) | 1.2 (0.5) |
| *Missing* |  | 2 (29%) |  |  |
| REM (minutes) | 65.1 (32.8) | 48.2 (13.9) | 61.5 (23.5) | 17.1 (17.6) |
| *Missing* |  |  | 1 (10%) | 2 (18%) |
| NRH (%) | 4.9 (4.1) | 5.9 (4.2) | 3.8 (4.4) | 13.9 (10.7) |
| *Missing* |  |  | 1 (10%) | 2 (18%) |
| Sleep efficiency (%) | 77.7 (9.0) | 62.1 (15.7) | 75.4 (10.0) | 69.8 (16.6) |
| *Missing* |  |  | 1 (10%) | 2 (18%) |
| AAI (events/hour) | 13.0 (8.2) | 22.0 (15.8) | 43.7 (27.3) | 14.5 (12.0) |
| *Missing* | 1 (11%) | 1 (14%) | 2 (20%) | 2 (18%) |
| Spindle duration (minutes) | 1.7 (1.9) | 0.6 (0.8) | 6.0 (14.0) | 5.3 (12.6) |
| *Missing* |  |  | *1 (10%)* | *2 (18%)* |
| AN3 (%) | 3.8 (3.4) | 1.8 (2.4) | 2.4 (5.0) | 5.4 (6.1) |
| *Missing* |  |  | *1 (10%)* | *2 (18%)* |

In the context of sleep, the DLB group exhibited the lowest amount of time spent in REM and the highest percentage of NRH. The control, MCI, and AD groups had similar REM and NRH sleep values to one another. Although the MCI group had the lowest sleep efficiency, the results were similar across all 4 groups. The number of autonomic activations per hour was highest in the AD group, whereas the other groups had lower values that were close to each other. Both the AD and DLB groups exhibited longer mean spindle durations with large variability, whereas the control and MCI groups exhibited shorter mean spindle durations with little variability. Finally, all four groups spent only a small percentage of sleep time in AN3 without any clear group differences.

## DISCUSSION AND IMPLICATIONS

To our knowledge, this preliminary study is among the first to perform both objective accelerometry and EEG-based sleep measures in older controls and in individuals with MCI, AD, and DLB. These preliminary analyses illustrate the promise of using accelerometry and at-home sleep devices as objective digital biomarkers for the early detection of dementia, as we found suggestive differences between groups, particularly when comparing MCI to dementia.

As expected, the cognitive performance of the MCI group was closest to that of controls, with delayed memory notable as the most affected domain, and the other domains slightly below the average performance of controls. Surprisingly, we also found that the MCI group had lower physical activity levels than the AD group and the worst sleep efficiency of any study group.

Other studies have found that individuals with MCI may be less physically active than controls^31^ and that PSG may be able to differentiate individuals with MCI from controls,^32^ but we would not necessarily expect participants with MCI to have lower sleep efficiency than individuals with dementia.^33^ It is possible that medication effects may have complicated these analyses or that participants with AD are experiencing shorter periods of sleep latency than participants with MCI and that these differences are then characterized as better sleep efficiency. In any case, these findings should be interpreted cautiously given the small sample size, but in a broader sense they point to activity and sleep disruption as changes in behavior that emerge prior to significant cognitive decline. Indeed, the contrast between cognitive performance and activity and/or sleep suggests that other lifestyle factors (e.g., physical health and mood) may affect daily activity and sleep before substantial cognitive decline is evident. In a general sense, these results align with longitudinal studies indicating that changes in physical activity are more pronounced several years prior to incident dementia and are most apparent at the time of diagnosis.^9,34^ Clinically, this emphasizes the importance of investigating both cognitive and physical characteristics when early stages of a neurodegenerative disorder is suspected.

It is also encouraging that in these preliminary findings, the accelerometry and sleep monitoring seemed to detect differences between the DLB group and other groups, including the AD group. As expected, we found that despite having the youngest mean age, the DLB group exhibited the highest UPDRS Part III scores (indicating more severe motor signs and symptoms) and NPI scores (indicating higher frequency of neuropsychiatric symptoms). These observations are not surprising, as individuals with DLB tend to experience a range of parkinsonian symptoms (e.g., bradykinesia, motor rigidity), neuropsychiatric symptoms (e.g., hallucinations or delusions), and sleep-related symptoms (e.g., RBD and daytime sleepiness),^24^ all of which may decrease or disrupt physical activity and sleep. Some of these symptoms can be captured by our digital biomarkers, as the DLB group exhibited the lowest physical activity levels, spent the least amount of time in REM sleep, and spent the highest percentage of time in NRH (a novel sleep biomarker). Moreover, the REM- and NRH-related findings are consistent with previous studies that showed that the accumulation of α-synuclein pathology within cholinergic neurons in the brain contributes to disrupted REM sleep regulation in synucleinopathies like DLB.^35^

Likewise, the AD group had levels of cognitive impairment that were similar to the DLB group, but as expected, the AD group performed better in the visuospatial and attention domains. This finding aligns well with the established literature on domain-specific cognitive impairments for AD and DLB.^36–38^ Interestingly, the AD group also had activity counts, REM sleep levels, and NRH levels that were intermediate between the DLB and control groups and, in some cases, closer to the MCI group. This kind of intermediate physical activity has been reported by Gramkow et al.^39^ and may be related to cognitive decline rather than overt motor symptoms.

We also found that the AD group had the highest and most variable novel sleep biomarker AAI in the study and more efficient sleep and longer sleep spindle duration than the MCI or DLB groups; the latter finding is surprising, as we would have expected the AD group to fall between the MCI and DLB groups in sleep efficiency and spindle duration.^33,40,41^ These unexpected sleep results may be attributable to a combination of factors, including, perhaps, (1) the heterogeneity of AD and DLB (e.g., variability in disease onset, progression, and underlying pathophysiology); (2) the unknown effects of medications (i.e., medications that are known to interfere with sleep architecture were excluded) or comorbid medical conditions; and (3) the presence of outliers in our small sample.

Finally, as expected, healthy controls had the lowest UPDRS Part III scores, the highest physical activity counts, and the best sleep efficiency. Although the spindle duration finding named above was unusual, the other findings are consistent with the known connection between sleep quality and preserved cognition.^42,43^

As we have suggested, a major limitation of the study is the small sample size, which limited our analyses to descriptive statistics, prevented us from analyzing MCI as prodromal AD and DLB subtypes, and contributed to analytic challenges related to outliers and missing data. However, it is reassuring that the clinical characteristics of each group (as measured by the NPI and MoCA) were similar to the reports in the literature. Future studies should enroll a larger cohort to improve generalizability and expand upon the relationships between physical activity, sleep, cognition, and disease type.

A key strength of this study was our use of objective, continuous measures of both sleep and physical activity. Our study was able to observe millions of daily activity counts per participant rather than needing to qualitatively categorize participants into low-, medium-, or high-activity groups. This meant we were able to quantitatively capture all levels of physical activity, including low-intensity, incidental activities that may be more characteristic of older adults. In addition, many accelerometry studies have a shorter wear time than we used in this study; our participants wore wrist monitors over 14 consecutive days, and thus, we were able to observe habitual activity patterns that may be better at differentiating between control, MCI, and dementia populations.^19,44^ Moreover, although our overall sleep efficiency and microarchitectural findings showed some inconsistencies compared to previous studies, our use of objective sleep monitoring technology allowed us to examine sleep architecture in participants with a range of cognitive abilities, including participants with MCI, AD, or DLB. Future studies should continue to leverage these technologies to investigate prodromal manifestations of dementia and to perform longitudinal follow-ups that examine how alterations in activity and sleep microarchitecture may affect health outcomes.

## CONCLUSION

This study is among the first to deploy an observational approach to objectively assess physical activity and sleep in control, MCI, AD, and DLB groups, integrating traditional cognitive and motor assessments, accelerometry, and PSG/EEG-based sleep assessments. Our preliminary findings help characterize the profiles of physical activity, sleep, cognition, and behavior in these groups and may be useful for identifying early behavioral markers of neurodegenerative disorders. Future work should incorporate larger samples and longitudinal follow-ups when utilizing accelerometers and Sleep Profiler to assess the role of physical activity and sleep on cognitive performance.

## Funding

This work was supported by the National Institute on Aging (R21/R33 AG064271 to DWT) and was supported in part by the U.S. Department of Veterans Affairs Office of Research and Development.

## Conflicting Interests

None reported.

## Data Availability

Data used in this study is available upon request, subject to VA regulations and with corresponding author’s approval.

## Author Contributions/CRediT

Conceptualization: ASD and DWT; data curation: DL, ES, SP, VLO, and YC; formal analysis: KB and KW; funding acquisitions: ASD and DWT; investigation: DWT, SCH, SP, and VLO; methodology: DWT, PD, and SP; project administration: ASD, DWT, SP, and VLO; supervision: DWT, PD, and SCH; writing—original draft: ASD, KB, YC, and VLO; writing— review and editing: ASD, DL, DWT, ES, KB, KW, PD, SCH, SP, VLO, and YC.

## Acknowledgments

The authors would like to thank all participants and companions for their invaluable contributions that made this research possible.

## Notes

### Competing Interest Statement

The authors have declared no competing interest.

### Author Declarations

VA Puget Sound Health Care System Institutional Review Board #1 gave ethical approval for this work.

